# Early antiparasitic treatment prevents progression of Chagas disease: Results of a long-term cardiological follow-up study in a pediatric population

**DOI:** 10.1101/2020.06.30.20143370

**Authors:** Nicolás González, Samanta Moroni, Guillermo Moscatelli, Griselda Ballering, Laura Jurado, Nicolás Falk, Andrés Bochoeyer, Alejandro Goldsman, María Grippo, Héctor Freilij, Eric Chatelain, Jaime Altcheh

## Abstract

**Background:** Chagas disease (CD), a parasitic disease caused by *Trypanosoma cruzi*. Parasite persistence is crucial in the development and progression of Chagas cardiomyopathy that occurs in 30% of untreated patients.

**Methods and findings:** A cohort of 95 CD treated children, with at least 6 years post-treatment follow up, was evaluated. Median after treatmeant follow-up was 10 years. At the time of the last visit a group of non infected subjects were also included as a control for cardiological studies.

During follow-up, the majority of treated subjects 59/61 (96%) achieved negative parasitemia by qPCR at the end of treatment. A decrease in *T. cruzi* antibodies titers were observed and seroconversion by two conventional serology tests (IHA, ELISA) occurred in 53/95 (56%).

Holter showed alterations in 3/95 (3%) of treated patients: isolated ventricular extrasystoles and nocturnal sinus bradycardia (one patient); asymptomatic and vagal related 1st and 2nd degree AV block (one patient); and complete right bundle branch block (cRBBB) (one patient). Only the last one is probably related to CD involvement. 2D speckle tracking echocardiography was conducted in 79/95 (83%) patients and no alterations in myocardial contractility were observed.

In the non infected group Holter evaluations showed similar non pathological results in 3/28 (10%) of subjects: isolated ventricular premature beats (2 patients); asymptomatic 2nd degree AV block with Wenckebach sequences during night time (one patient). 2D speckle tracking echocardiography was conducted in 25/28 (89%) with no alterations.

**Conclusions:** After long term follow-up of a cohort of treated children for CD, a good parasiticidal treatment effect and a low incidence of cardiological lesions, related to Chagas disease, were observed. These results suggest a protective effect of treatment on the development of cardiological lesions and strengthen the recommendation of early diagnosis and treatment of infected children.

**Trial registration:** ClinicalTrials.gov NCT04090489.

**Author summary:** If left untreated, CD evolves into a chronic oligosymptomatic infection that can progress to cardiac complications in 30% of patients after several years.

It is known that the main early marker of cardiac involvement are alterations in the conduction system. There are few studies of long-term after treatment cardiological evolution which have assessed the clinical effectiveness of treatment.

The rationale for CD treatment is to avoid the development of cardiological complications. The parasiticidal effect of treatment has been demonstrated but its clinical effectiveness in preventing cardiac involvement requires long term follow-up.

In our long term follow-up study of treated children, we observed the preventive effect on cardiac lesions by treatment with benznidazole or nifurtimox. This intensifies the need for early diagnosis and treatment to prevent the development of long term complications observed in CD.

## Introduction

Chagas disease (CD), caused by *Trypanosoma cruzi (T. cruzi)*, has a broad range of hosts, and is primarily transmitted through infected triatominae bugs or congenitally. [1] Historically considered a regional disease, CD has now become a global phenomenon due to migration, reaching regions such as Europe, USA, Australia and Japan. [2-5]

CD presents with an acute phase with high parasitaemia that, if left untreated, evolves into a chronic oligosymptomatic infection that can progress in 30% [6] of patients to cardiac and/or gastrointestinal complications after several years.

Electrocardiogram (ECG) alterations are early signs of cardiac involvement. Some ECG hallmarks of CD are cRBBB, QRS complex widening or fragmentation, and sinus bradycardia. In more advanced stages, direct damage to the cardiac conduction system is revealed by severe cardiac alterations such as second and third-degree AV block, atrial arrhythmias, ventricular premature beats or ventricular tachycardia. [6-7]

Myocardial deformation imaging is a new technique for quantitative assessment of myocardial contractility. The novel method of speckle-tracking 2D echocardiogram (STE) for myocardial strain imaging can quantitatively assess myocardial contractility deformation, and has been validated for the detection of subclinical left ventricular dysfunction as an early marker of cardiac compromise in CD patients [8]. STE can play an important role in the evaluation of CD patients, especially in the early chronic stage when segmental ventricular wall motion abnormalities are more subtle.

Parasite persistence is crucial to the development and progression of CD cardiomyopathy [9]. Effective antiparasitic treatment of CD could, therefore, prevent complications related to the disease by eliminating tissue parasites. However, there is insufficient data about the impact of medication on the prevention of cardiac lesions, since this requires long-term follow-up of treated patients.

The aim of this study is to describe the incidence of cardiological alterations, the kinetics of *T. cruzi* antibodies and treatment response using *T. cruzi* - PCR after long-term follow-up of a cohort of treated CD children.

## Methods and materials

A prospective cohort study of pediatric CD patients treated with benznidazole (Bz) or nifurtimox (Nf) with long-term follow-up was conducted at the Servicio de Parasitología y Chagas, Hospital de Niños Ricardo Gutiérrez, a tertiary referral center in Buenos Aires, Argentina. Children with at least 6 years post-treatment follow-up and who attended a clinical visit between August 2015 and November 2019 were considered for participation in the study.

In order to evaluate the prevalence of normal ECG findings in a group with similar ethnic and socioeconomic characteristics, family members of the same age as treated cases, but without CD, were enrolled as cardiological controls.

### Inclusion criteria

- Children with CD treated with Bz or Nf with at least 6 years of post-treatment follow-up. In infants younger than 8 months CD was diagnosed by direct observation of *T. cruzi* using a parasitological concentration method (microhematocrit test - MH) or xenodiagnosis, and in infants older than 9 months, by two reactive serological tests (ELISA, and indirect hemagglutination - IHA).
- Signed informed consent or assent according to patient age.

### Exclusion criteria

- Patients with chronic diseases (renal, hepatic, neurological) that could affect the interpretation of the results (at the discretion of the researcher).
- Subjects with congenital heart disease.

### Treatment

Bz (5-8 mg/kg in 2 or 3 doses) and/or Nf (10-12 mg/kg in 2 or 3 doses) was prescribed for 60-90 days. Medication was provided in monthly batches and compliance was assessed by counting the remaining tablets at each visit.

### *T. cruzi* serology and qPCR

Blood samples were collected at diagnosis, end-of-treatment and every 6-12 months after thereafter. Detection of *T. cruzi*-DNA by PCR was carried out in blood (2 mL) mixed with EDTA-guanidine buffer. [10] Serology tests were performed by IHA and ELISA.

### Cardiological evaluation

ECG was performed at diagnosis and every year following treatment. In addition, cardiological evaluation using 24-hour Holter monitoring and STE were performed at the time of the final study follow-up visit. Cardiological studies were carried out at the Cardiology service of the Hospital de Niños Ricardo Gutiérrez, Buenos Aires, Argentina by trained pediatric cardiologists and pediatric electrophysiologists.

### Ethics

The study was approved by the ethics committee of the Hospital de Niños Ricardo Gutiérrez, Buenos Aires, Argentina.

Written informed consent by the patient and healthy controls and/or parents was obtained for all patients according to age and local regulations.

The protocol was registered with ClinicalTrials.gov, NCT04090489.

### Statistical analysis

Continuous variables are presented as means with CI95% or medians and interquartile range, and categorical variables as percentages.

The disappearance kinetics of *T. cruzi* serum antibodies were analyzed using survival analysis. Analyses were performed with R software v3.0 (R Core Team 2018, R Foundation for Statistical Computing, Vienna, Austria https://www.R-project.org/).

## Results

A total of 95 patients (40 male, 55 female) previously treated for CD were enrolled, as well as 28 uninfected children as matched controls for the ECG evaluation (Fig 1). In CD patients, the median age at diagnosis was 4.28 years (range 10 days −20 years); median age at final evaluation was 15 years (range 6-33 years). Subjects were all born in Argentina, and mainly infected through congenital transmission. Following treatment with Bz or Nf, all enrolled subjects continued to reside in Buenos Aires or the Greater Buenos Aires area, where there is no vector transmission that could cause reinfection.

**Fig 1.**
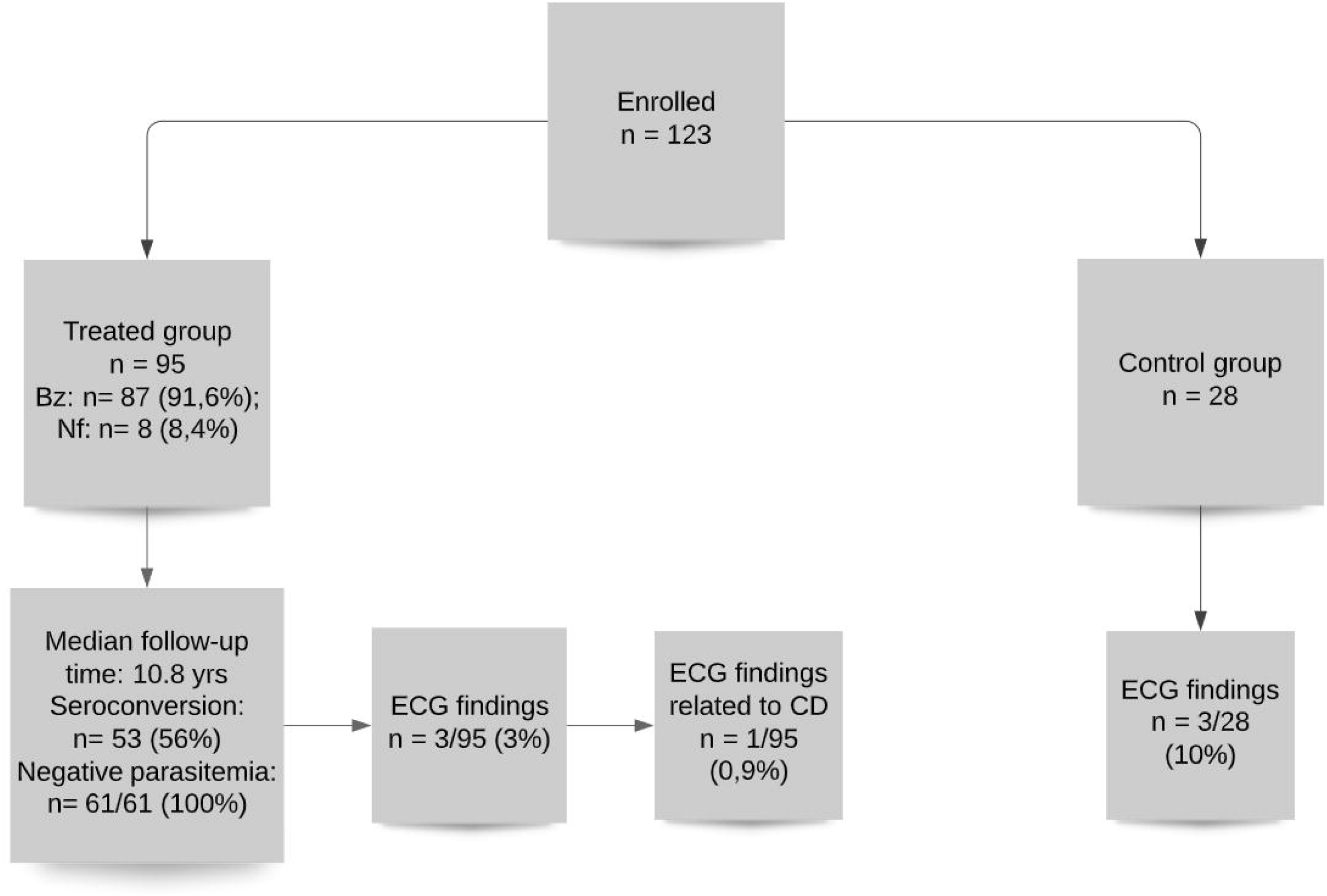
Flow chart of patient enrollment in the study.

The median age of the non-infected control group was 13.2 years (range 6-25 years). These subjects were family members of the infected cases.

The parasiticidal treatments prescribed were Bz in 87 patients and Nf in 8 patients. In 6 cases, treatment was prescribed more than once (see below). Based on tablet counts and medication log reviews, good compliance both for Bz and Nf treatment appears to have been achieved.

Mean Bz dose was 6.5 mg/kg per day (range 5–9.2 mg/day) divided in two (n=84) or three doses (n=3). In 3 cases, data on daily frequency intake were not available. Mean treatment length was 59.8 days (30–60). A total of 80/87 (91.9%) enrolled patients completed drug treatment as prescribed. Bz was well tolerated and adverse drug reactions (ADRs) were mild, requiring treatment suspension in only 4 (4.5%) patients. Of the three remaining patients, two received 30 and 38 days of treatment respectively, while the third received a new cycle of treatment (see below).

Mean Nf dose was 10.8 mg/kg per day (range 5.7–13.9 mg/kg/day) divided in two (n=7) or 3 doses (n=1). Median treatment length was 69.5 days (55–90). All enrolled patients completed drug treatment as prescribed. Nf was well tolerated and ADRs were mild, not requiring treatment suspension.

Six patients received more than one course of treatment: one patient underwent treatment with Bz and, due to difficulties in adherence, a second course of Bz was administered to complete 60 days. Two patients completed their course of treatment with Bz (at the age of 3 and 6 years, respectively), but due to persistently positive qPCRs after treatment, they were considered as treatment failures, and a new course of treatment was indicated; in one case, treatment failure was attributed to a lack of compliance with Bz treatment and so a new course of Bz was prescribed. For the second patient, where there was evidence of good compliance to Bz, the outcome was assumed to be true treatment failure and a course of Nf was prescribed. Both patients showed good treatment compliance and tolerance, and good treatment response with a consistent decrease in *T. cruzi* antibodies and negative qPCR after treatment. In three patients, Bz treatment was suspended due to skin adverse events and all of them subsequently received a new course of treatment with Nf. All three patients showed good tolerance and good treatment response.

Baseline parasitemia data were available for 65 patients: 61 (93%) were initially positive (46 by qPCR, 6 by both qPCR and MH, 7 by MH and 2 by xenodiagnosis). The majority of treated subjects 59/61 (96%) achieved negative parasitemia by *T. cruzi* -qPCR at the end of treatment and remained negative throughout the follow-up period.

In two cases, qPCR results remained positive after treatment; these two patients were considered as treatment failures and a new course of treatment was prescribed, with good response (see above).

Reduction in *T. cruzi* antibody titers assessed by conventional serology (IHA and ELISA) was observed during after treatment follow-up in all treated patients. IHA tests became negative in 72/95 (76%) at a median time of 3.17 years (95%CI: 2.38 − 5.76) and ELISA tests became negative in 57/95 (60%) of patients at a median time of 4.68 years (95%CI: 2.87 − 14.50). In 53/95 (56%) both serological tests became negative at a median time of 6 years (95%CI: 4,6 - infinity) (Fig 2).

**Fig 2.**
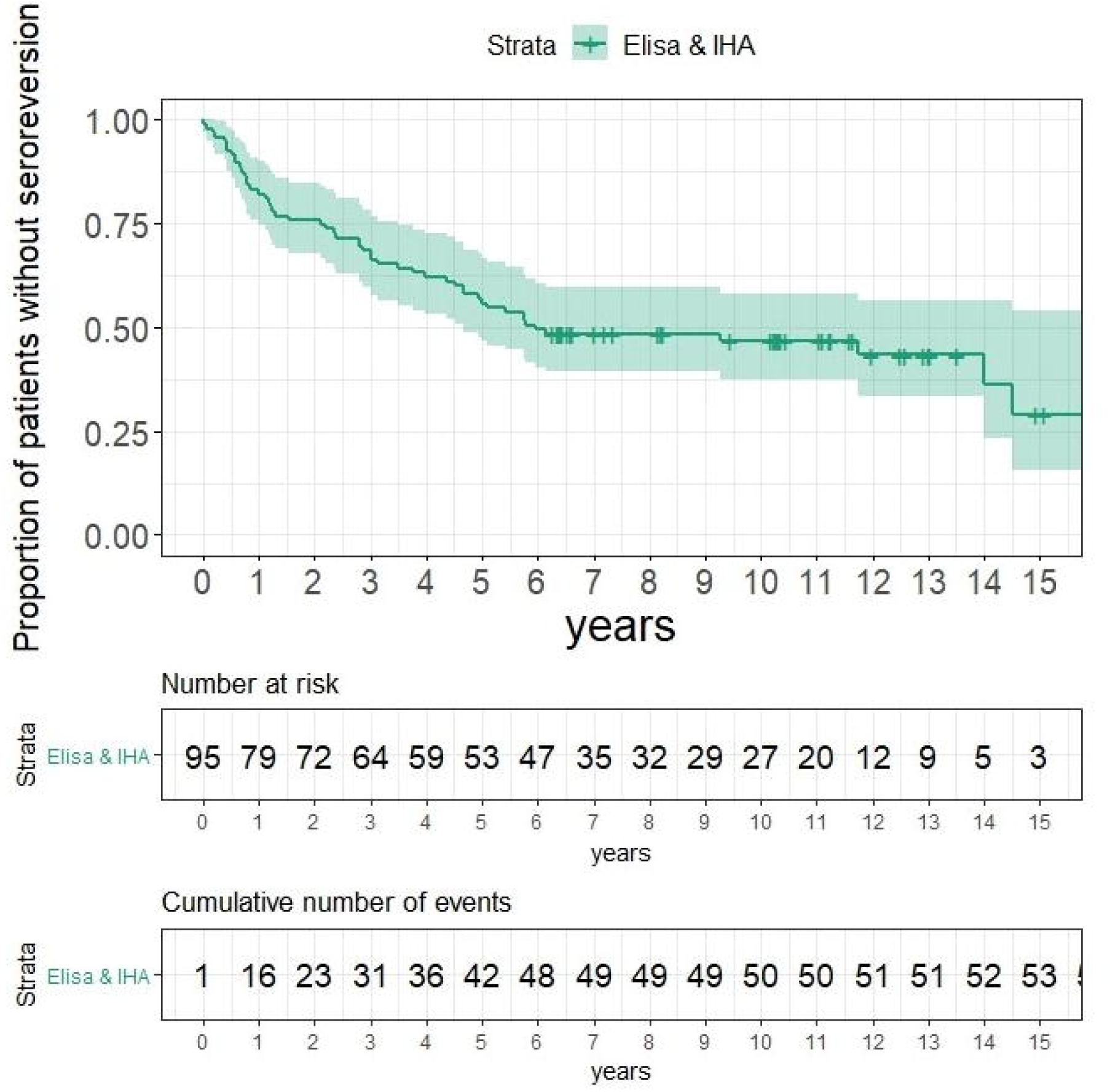
Serological seroconversion by ELISA and IHA profile in treated patients (Kaplan-Meier).

### Cardiological evaluation

24 hour Holter monitoring and STE studies were conducted in both groups, treated patients and non-infected controls. In the treated group, evaluation by Holter was carried out at a median of 10 years after treatment (range 6 − 27 years). This evaluation showed findings in 3/95 (3%) of treated patients (Table 1), including isolated ventricular extrasystoles and nocturnal sinus bradycardia (one patient); asymptomatic and vagal related 1st and 2nd degree AV block (one patient); and cRBBB (one patient). STE was conducted in 79/95 (83%) of patients, including the subjects with Holter alterations. No alterations in myocardial contractility were observed in any patients.

**Table 1.** Cardiological findings.

| Treated patients group |  |  |  |  |  |  |  |
| --- | --- | --- | --- | --- | --- | --- | --- |
| Patient ID | Age at treatment (yrs) | Age at evaluation (yrs) | Treatment | Serology at evaluation | STE | Holter finding | Related to CD |
| ID 24 | 11 | 22 | Bz 60 days<br>Good compliance | ELISA: Reactive<br>3.3<br>IHA: Negative | Normal | Isolated ventricular extrasystoles and nocturnal sinus bradycardia | No |
| ID 41 | 1 | 11 | Bz 60 days<br>Good compliance | Negative | Normal | Asymptomatic and vagal related<br>1st and 2nd degree AV block | No |
| ID 70 | 3 | 19 | Bz 70 days<br>Good compliance | Negative | Normal | Complete right bundle branch block | Probable |

| Non- infected matched control Group |  |  |  |
| --- | --- | --- | --- |
| Patient ID | Age at evaluation (yrs) | STE | Holter finding |
| ID 8 | 13 | Normal | Asymptomatic 2nd degree AV block with Wenckebach sequences during the night |
| ID 18 | 10 | Normal | Isolated ventricular premature beats |
| ID 27 | 9 | Normal | Isolated ventricular premature beats |

In the non-infected group, Holter evaluations yielded similar results to those in the treated group; 3/28 (10%) of subjects had non-specific ECG findings (Table 1), including isolated ventricular premature beats in 2 patients and asymptomatic 2nd degree AV block with Wenckebach sequences during the night in one patient. STE was conducted in 25/28 (89%) of patients and no alterations in myocardial contractility were observed in any of the cases evaluated.

## Discussion

Currently, CD treatment with Bz or Nf is recommended as the standard of care for acute and early chronic phases in children. In the early phase of infection, which is more easily observed in children, the majority of cases are asymptomatic and without cardiac involvement, as described in this study. However, approximately 30% of untreated patients with CD will eventually develop cardiomyopathy in the chronic stage. [7, 11] Multiple mechanisms may be responsible for the development of cardiac lesions in T. cruzi infection. The presence and persistence of the parasite is a critical factor which triggers a specific immune response, inducing vascular endothelial cell damage. This may play an important role in the pathogenesis of CD cardiac involvement. [12]

At tissue level, a diffuse cellular infiltrate with microcirculation alterations affects the parasympathetic innervations, increasing sympathetic activity, and leading to further cardiac damage and development of arrhythmia. [13]

The treatment of infected subjects is, therefore, based on the elimination of intracellular parasites to avoid the development of future cardiological complications.

In previous studies [14], we described the effectiveness of parasiticidal treatment in children. There was a decrease in T.cruzi antibodies measured by conventional serology and persistent non-detection of T.cruzi-qPCR during after-treatment follow-up. The parasiticidal effect of Bz and Nf was demonstrated by the clearance of parasitemia.

Electrocardiographic abnormalities are often the first indicator of CD cardiac involvement. The earliest ECG abnormalities of Chagas cardiomyopathy usually involve abnormalities of the conduction system, manifesting most frequently as different degrees of sinus dysfunction, right bundle branch block and / or left anterior fascicular block. Second and third degree atrioventricular blocks, as well as ventricular arrhythmias, have also been described in relation to advanced CD lesions.

There are few studies that have investigated the clinical effectiveness of CD treatment through the evaluation of cardiological events by long-term follow-up of asymptomatic treated children. It is important to note that children are different from adults. Although the basic principles of cardiac conduction and depolarization are the same as for adults, age-related changes in the anatomy and physiology of infants and children produce normal ranges for electrocardiographic features that differ from adults and vary with age. Conduction intervals (PR interval, QRS duration) are shorter than adults due to smaller cardiac size. First degree AV block and Wenckebach sequence may be a normal finding in healthy children during rest or under vagal tone. The QT interval depends on heart rate and age. Heart rates are much faster in neonates and infants, decreasing as the child grows older. Thus, cardiological evaluation in children must be conducted by pediatric cardiologists since features that would be diagnosed as abnormal in adult’s ECG may be normal, age-related findings in a pediatric ECG trace. This can be explained by the way the heart develops during infancy and childhood. The right ventricular dominance of the neonate and infant is gradually replaced by left ventricular dominance so that by 3-4 years the pediatric ECG resembles that of adults. In addition, some pathological findings could be related to technical issues and in other cases may be related to congenital lesions and not produced by T. cruzi infection. Awareness of these differences is the key to correct interpretation of pediatric electrocardiograms. [15]

In non-treated CD patients, the overall reported rate of cardiac disease progression is between 13 and 25 % [16] with a 2 to 4 % annual progression rate. [17] In our cohort, the median after-treatment follow-up time was 10 years (6.02 − 26.9) and only one patient showed ECG alterations that could possibly be related to T. cruzi infection. The other ECG findings observed (which could be considered abnormal from an adult ECG perspective) were in fact likely to be normal ECG findings in our pediatric cohort of patients, and therefore unrelated to CD, with only one patient showing signs compatible with cardiac involvement in CD (i.e. complete right bundle branch block) [7]. The other two patients with Holter findings showed non-specific ECG alterations that are currently considered normal variations. To correct for the limited amount of data in healthy populations with similar ethnic and socioeconomic characteristics as our patients, we enrolled a non-infected group of volunteers as a control group in order to better establish a “normal” baseline for the Holter evaluation. Notably, no differences in the prevalence of ECG normal variations were found between the treated and uninfected groups.

Several published studies which reported ECG abnormalities in children with CD showed alterations that are unlikely to be related to CD, and in many cases should simply be considered as normal variants for age. [18-20] In a 3-year follow-up of treated children in the chronic phase, it was shown that only one child in the Bz group and four in the control group developed complete right bundle branch block (p: 0.36). At the 6-year follow-up time-point, no progression of cardiac lesions was observed.

In another study, 18.6% of treated children showed ECG changes during follow-up. Only four children presented alterations of right bundle conduction that could be attributed to CD but which were not pathognomonic lesions due to CD. The risk of incidence of ECG alterations comparing treated versus untreated children was 0.74 (95% CI: 0.28 −1.97, p: 0.54), showing no differences between the group. [21] This study suggested that the treatment did not stop the appearance of new cardiac lesions. However, the anomalies described were mostly normal findings characteristic in children (i.e. extrasystoles, incomplete right bundle branch block, first-degree atrioventricular block) and unlikely to be related to CD.

Several observational studies in adults have shown the positive effect of treatment on the prevention of the development of cardiac alterations. [22-25]

In contrast, in the BENEFIT study [26] no difference between treated adult subjects and the placebo group was observed. However, this was a different population in which severe cardiac pathology was already present and by this stage the cardiological damage was probably irreversible and not modifiable by the parasiticidal treatment.

In our study, all evaluated patients showed normal myocardial contractility as measured by STE. Over the last few years echocardiography has become the most common imaging modality used to assess and follow-up patients with CD. In the early stages of cardiac involvement, ventricular wall motion abnormalities are observed, and STE allows a more precise measurement of regional myocardial function. [8]

The results of the current study confirmed the efficacy and safety of Bz and Nf in pediatric CD patients. In addition, we did not find a higher incidence of ECG alterations in patients treated in childhood for CD with a long-term after-treatment follow-up, which could be related to a protective effect of treatment at an early stage of T. cruzi infection.

Presently, the standard of care for CD is the universal treatment of all infected children. The results of this study support early screening for T. cruzi infection in children and infants from infected mothers so that timely treatment can be provided. Early diagnosis and treatment could prevent or slow the progression of cardiac involvement in chronic CD.

We concluded that all treated patients showed parasitological and serological signs of treatment response (seroreversion in more than half of the patients, constant negative qPCR and a decrease in antibodies titers for the others). More importantly, we have shown that the development of cardiac alterations was prevented by parasiticidal treatment after a median follow-up of over a decade, apart from one patient who showed signs of potential cardiac pathology related to CD. These results further underline the importance of early diagnosis and availability of treatment, especially for children, young adults and women of childbearing age, in order to prevent new infections and development of symptomatic chronic Chagas disease. Further studies with long-term follow-up (20-30 years) of the present cohort will be conducted in order to confirm that early treatment prevents later cardiological compromise in CD patients.

## Data Availability

Individual participant data will be available (including data dictionaries).
All of the individual participant data collected during the trial, after de-identification, will be shared.
Study protocol, statistical analysis plan, informed consent form, clinical study report and analytic code will be available, too.
It will be available beginning 9 months and ending 36 months following article publication.
Information will be shared with investigators whose proposed use of the data has been approved by an independent review committee (learned intermediary) identified for this purpose.
It would be used for individual participant data meta-analysis.
After 36 months the data will be available in our service's data warehouse but without investigator support other than deposited metadata.

## Abbreviations

Bz: benznidazole
CD: Chagas disease
cRBBB: complete right bundle branch block
ECG: electrocardiogram
Nf: nifurtimox
STE: speckle-tracking 2D echocardiogram

## Supporting information

**S1 File. STROBE Statement**. Checklist of items that should be included in reports of cohort studies.

